# Radiobiological meta-analysis of the response of prostate cancer to high dose rate brachytherapy

**DOI:** 10.1101/2024.07.12.24310000

**Authors:** Eva G Kölmel, Miguel Pombar, Juan Pardo-Montero

**Author notes:** **Correspondence:** Juan Pardo-Montero, Grupo de Física Médica e Biomatemáticas, Instituto de Investigación Sanitaria de Santiago (IDIS), Servizo de Radiofísica e Protección Radiolóxica, Hospital Clínico Universitario de Santiago, Trav. Choupana s/n, 15706, Santiago de Compostela (Spain).

## Abstract

**Background and purpose:** Clinical studies have shown a marked reduction in tumor control in prostate cancer treated with radically hypofractionated high-dose-rate brachytherapy (HDR-BT). The purpose of this study was to analyze the dose-response of prostate cancer treated with HDR-BT, specifically aiming at investigating the potential failure of the linear-quadratic (LQ) model to describe the response at large dosesper-fraction.

**Materials and methods:** We collated a dataset of dose-response to HDR-BT (3258 patients). The analysis was conducted separately for low and intermediate risk, resulting in 21 schedules (1643 patients) and 22 schedules (1615 patients), respectively. Data were fitted to tumor control probability models based on the LQ model, the linear-quadratic-linear (LQL), and a modification of the LQ model to include the effect of reoxygenation during treatment.

**Results:** The LQ cannot fit the data unless the α/β is allowed to be very high (∼[20-100] Gy, 95% confidence interval). If the α/β is constrained to be low (< 8 Gy) the LQ model cannot reproduce the clinical results, and the LQL model, which includes a moderation of radiation damage with increasing dose, significantly improves the fitting. On the other hand, the reoxygenation model does not match the results obtained with the LQL.

**Conclusion:** The clinically observed reduction in tumor control in prostate cancer treated with radical HDR-BT is better described by the LQL model. Using the best-fitting parameters, the BED for a 20 Gy × 1 treatment (95 Gy) is far less than that of a conventional 2 Gy × 37 fractionation (184 Gy). These results may assist in the design of radical HDR-BT treatment.

## Introduction

Radiotherapy is widely used to treat prostate cancer. Radiotherapy options include external beam radiotherapy **[1]**, proton therapy **[2, 3]**, and brachytherapy, either low-dose-rate or high-dose-rate (HDR-BT) **[4, 5]**. HDR-BT is nowadays used as monotherapy for many patients **[6]**, achieving good clinical outcomes, and because it is delivered with hypofractionated schedules, reducing treatment time and increasing patient comfort **[5]**.

The response of prostate cancer to radiotherapy has been extensively studied **[7, 8, 9]**. The consensus is that the *α*/*β* ratio of prostate cancer is low (typically in the 1-4 Gy range), and therefore this tumor is very sensitive to fractionation. Nonetheless, alternatives to the low *α*/*β* have been suggested, like tumor hypoxia **[10]**. In recent years, Stereotactic Body RadioTherapy (SBRT) has become widely used to treat prostate cancer **[11]**, with doses per fraction reaching up to 10 Gy. The response of prostate cancer to hypofractionated SBRT protocols has been recently analyzed **[12, 13, 14]**. All three studies reported low *α*/*β* ratios, in agreement with values obtained from lower doses per fraction. HDR-BT is delivered with hypofractionated protocols that are even more radical, reaching >20 Gy in a single fraction. However, several HDR-BT clinical trials have shown a marked reduction in tumor control (<70%) when delivering single fraction treatments with >20 Gy. This loss in tumor control is not supported by a low *α*/*β* ratio. Guirado *et al*. have recently analyzed the response of prostate cancer to HDR-BT, suggesting a large *α*/*β* ratio (∼23 Gy) to explain the poor control achieved with HDR-BT single-fraction treatments **[15]**. They also argued that the linear-quadratic (LQ) model may not be adequate to describe the response to very large doses per fraction.

The validity of the LQ model for large doses per fraction has long been questioned **[16]**, with different studies suggesting either a moderation or a boost of the cell killing effect predicted by the LQ with increasing dose per fraction **[17, 18]**. The moderation of the cell killing effect predicted by the LQ with increasing doses might explain the clinical results obtained with HDR-BT single-fraction treatments. This effect can be modeled with the linear-quadratic-linear (LQL) **[19]**. In fact, some evidence of a LQL-like response in the dose-response curves of prostate cancer treated with external radiotherapy was recently discussed in Refs. **[13, 14]**.

The poor control obtained with HDR-BT single-fraction treatments could also be rationally explained because of hypoxia and reoxygenation, as originally suggested by Nahum *et al*. **[10]**. If tumors are hypoxic and reoxygenate during treatment, short protocols delivering larger doses per fraction may be suboptimal.

In this work we have collated and analyzed a dataset of dose-response for HDR-BT of prostate cancer. We have performed a radiobiological analysis of the dose-response, considering not only the LQ model but also more advanced models including damage saturation at large doses and re-oxygenation, aiming at advancing the understanding of the response of prostate cancer to very large doses per fraction.

## Materials and methods

### Clinical dataset

The clinical dataset was created following a two-step process: initially, we expanded upon previously compiled datasets reported in **[5, 20, 6]**; subsequently, we performed a systematic search in Pubmed (in July 2023) for articles published after 2018 (the publication year of **[5]**). From each study, we extracted the number of patients, the distribution of patients according to the risk level, the number/percentage of patients receiving androgen deprivation therapy (ADT), the dose per fraction, the total dose, the overall treatment time and schedule details (fractions per day, time intervals between fractions), and the 5 year control rate, with control defined as freedom from clinical or biochemical failure (PSA nadir + 2 ng/mL). Studies that did not report any of these variables were excluded. Some studies included slightly different fractionations, and in those cases, the most commonly used fractionation was included. If different studies reported on the same (or similar) cohort with different follow-up, only the most recent publication was considered.

Overall, the collated dataset contained data from 19 studies (3258 patients). Our analysis was conducted separately for low risk (IR) and intermediate risk (IR), resulting in 21 schedules (1643 patients) for LR and 22 schedules (1615 patients) for IR. A limited number of data were found for high risk, but they were ignored because the number of schedules was not large enough to conduct the analysis. Some studies stratified patients in more than three groups (e.g. “favorable intermediate risk”, “unfavorable intermediate risk” and “very low risk”). In such cases, these results were merged into a single group. The percentage of patients receiving ADT was included in the dataset, even though this variable was not used in this analysis.

An overview of the schedules included in the analysis is presented in Table 1, with further detailed information is presented in Supplementary Table 1.

**Table 1:** Overview of the characteristics of the schedules included in the analysis.

| Risk | Number of schedules | Total number of patients | Number of patients per schedule (range) | Dose per fraction (range) | Total dose (range) | Overall treatment time (range) | Control at 5 years (range) | References |
| --- | --- | --- | --- | --- | --- | --- | --- | --- |
| LR | 21 | 1643 | 2 – 288 | 6.0 – 20.5 Gy | 19 – 54 Gy | 0 – 27 days | 0.66 – 1.00 | [21–35] |
| IR | 22 | 1615 | 16 – 284 | 6.0 – 20.5 Gy | 19 – 54 Gy | 0 – 41 days | 0.63 – 0.996 | [21, 23, 25–30, 32, 33, 36–39] |

## Radiobiological models

We used three different models to investigate the dose-response of prostate cancer to HDR-BT:

1. The LQ model: the surviving fraction of tumor cells after a dose *d* is **[40]**,

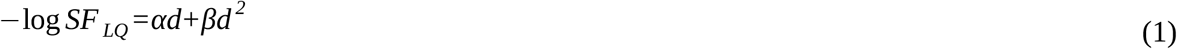

with *α* and *β* being the linear and quadratic parameters of the LQ model.
2. The LQL model: this model **[19]** includes a moderation of the quadratic term of the LQ model with increasing dose, which is controlled by the parameter *δ*:

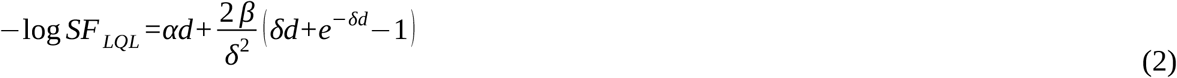
3. Stavrev’s model of reoxygenation: this model **[41]** relies on the LQ model, but with time-dependent *α* and *β* parameters to account for reoxygenation during treatment:

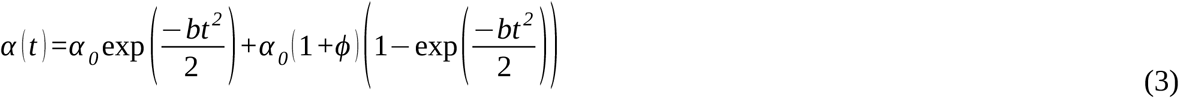

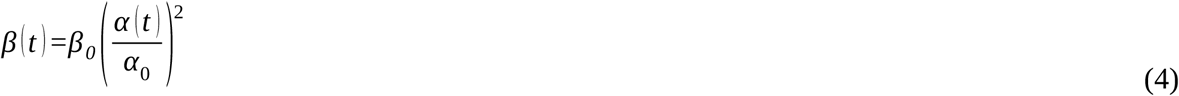

with parameters *b* and *φ* controlling the evolution of *α(t)* and *β(t)*. This results in a time dependent *α*/*β* ratio, which due to the quadratic dependence of *β* increases with time.

### Incomplete repair

Several schedules in the dataset delivered multiple fractions per day. In this situation, incomplete repair between consecutive fractions may play a role in the response to treatment. Therefore, we also investigated the possible contribution of incomplete repair by including it in the modeling. We used the LQ with incomplete repair correction **[42, 40]**. The surviving fraction of cells following the i-th radiation fraction is given by:

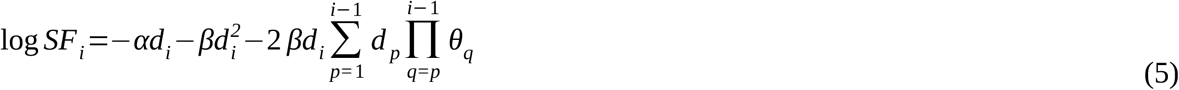

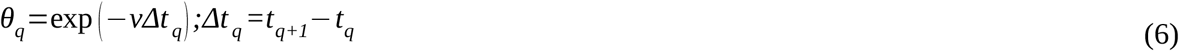

where *ν* is the repair rate of sub-lethal damage (we will refer instead to the half-life of damage, defined as *T*_*repair*_ = log 2/*ν*).

Incomplete repair was also considered in the LQL and reoxygenation models, using an expression identical to Eq. (5), but with *α(t*_*i*_) and *β(t*_*i*_) replacing *α* and *β* for the reoxygenation model (where *t*_*i*_ is the delivery time of the i-th fraction), and with an effective *β*-term replacing *β* for the LQL model, which can be obtained from Eq. (2):

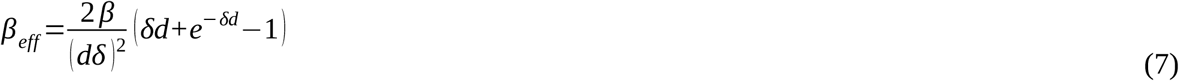

#### Overall surviving fraction and proliferation

When delivering *n* fractions in an overall treatment time *T*, the surviving fraction is given by:

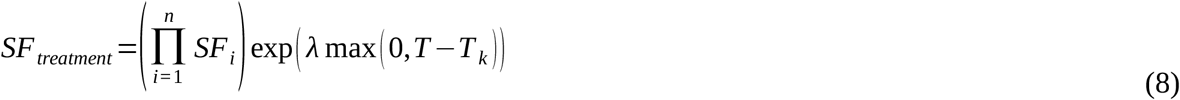

where *SF*_*i*_ is the surviving fraction associated to each fraction, and proliferation is modeled as exponential with rate *λ* after a kick-off time *T*_*k*_.

#### Tumor control probability and EQD2

The tumor control probability (*TCP*) was modeled using a logistic function **[43]**,

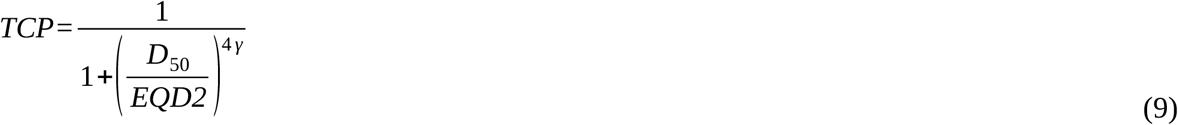

where *D*_50_ is the dose corresponding to 50% control (in 2 Gy fractions), and *γ* controls the slope of the dose-response curve. The *equivalent dose in 2 Gy fractions, EQD2*, of each schedule is model-dependent, and its explicit calculation for the models used in this work is shown in the Supplementary Materials.

### Statistical methods

We fitted the models to the clinical data with the maximum likelihood methodology, assuming binomial statistics for the reported control. We used an in-house developed algorithm **[14]** based on the simulated annealing method to perform the optimization (minimization of -log *L*, where *L* is the likelihood). Confidence intervals (CI) for the best-fitting parameters were obtained using the profile likelihood method **[44]**.

Fits with the LQ model have five free parameters (*α/β, λ’=λ/α, T*_*k*_, *γ, D*_*50*_). For the LQL model there is an extra free parameter, *δ*, and for Stavrev’s reoxygenation model there are two extra parameters, *φ* and *b*. When including incomplete repair there is an extra parameter for each model, *T*_*repair*_.

The space of parameter values was constrained to avoid reaching solutions that could be unphysical or not supported by biological data, and to speed up convergence. In particular, dose compensation due to accelerated proliferation was limited to *λ’* ≤ 2 Gy day^-1^, a limit well higher than the proliferation found in **[14]**, and the half-life of sublethal repair was limited to *T*_*repair*_ ≤ 6 h. For the *α*/*β* ratio, we employed two different constraints due to the discrepancies on the reported values from external radiotherapy and HDR-BT: on the one hand, a constraint 1 ≤ *α*/*β* ≤ 100 Gy to allow for large *α*/*β* ratios like those reported in **[15]**; on the other hand, a stronger constraint 1 ≤ *α*/*β* ≤ 8 Gy to force a low *α*/*β* ratio consistent with many reports from external radiotherapy.

The Akaike Information Criterion with sample size correction (AIC_c_) was used to evaluate the performance of the models **[45]**. The ΔAIC_c_ of a given *model* compared to the *reference* model (the LQ model in this work) is:

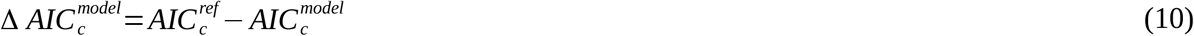

Models with lower AIC_c_ are preferred, *i*.*e*. positive ΔAIC_c_.

The implementation of the methodology was performed in Matlab (Mathworks, Natick, MA).

## Results

In Table 2, we present the best-fitting parameters and the goodness-of-fit (-log *L* and AIC_c_) obtained from fitting the LQ, LQL, and Stavrev’s reoxygenation models (with and without incomplete repair correction) to low and intermediate risk data. For these fits, the *α*/*β* ratio was allowed to lie in a large interval (1 ≤ *α*/*β* ≤ 100 Gy). In Figure 1 we present the TCP versus EQD2 curves (experimental data and model best-fits) obtained from this fitting strategy for the LQ, LQL, and reoxygenation models without incomplete repair correction. 95% confidence intervals for the *α*/*β* and *δ* (LQL) are presented in Supplementary Table 2.

**Table 2:** Best fits obtained with the LQ, LQL, and Stavrev’s reoxygenation model, LQ_ST_, without or with (subscript SD) sublethal damage incomplete repair correction, separated by risk (low, LR, and intermediate risk, IR). The table shows best fitting parameters, maximum likelihood, AIC_c_, and ΔAIC_c_ (referred to the LQ model) values. The symbol * indicates that the best-fitting parameter reached the edge of the constraint window.

| Risk | Model | Parameters |  |  |  |  |  |  |  |  |  |  |  |
| --- | --- | --- | --- | --- | --- | --- | --- | --- | --- | --- | --- | --- | --- |
| | | $\alpha/\beta$<br>[Gy] | $\lambda'$<br>[Gy day<sup>-1</sup>] | $T_k$<br>[day] | $\delta$<br>[Gy<sup>-1</sup>] | $\varphi$ | $b$<br>[h<sup>-2</sup>] | $T_{repair}$<br>[h] | $D_{50}$<br>[Gy] | $\gamma_{50}$ | $-\log(L)$ | AIC<sub>c</sub> | ΔAIC<sub>c</sub> |
| LR | LQ | 100* | 0.05 | 21.8 | - | - | - | - | 16.17 | 1.00 | 48.19 | 110.37 | - |
|  | LQL | 5.0 | 0.24 | 29.8 | 0.73 | - | - | - | 16.64 | 0.96 | 47.37 | 112.74 | -2.37 |
|  | LQ<sub>ST</sub> | 100* | 0.09 | 28.1 | - | 4.2E-2 | 2.5E-7 | - | 16.21 | 0.99 | 48.11 | 118.84 | -8.47 |
|  | LQ<sub>SD</sub> | 100* | 0.11 | 27.6 | - | - | - | 0.17 | 16.62 | 1.04 | 48.05 | 114.10 | -3.73 |
|  | LQL<sub>SD</sub> | 5.0 | 0.24 | 29.8 | 0.73 | - | - | 0.00* | 16.64 | 0.96 | 47.37 | 117.35 | -6.98 |
|  | LQ<sub>ST,SD</sub> | 100* | 0.11 | 27.6 | - | 0.00* | 0.00* | 0.17 | 16.62 | 1.04 | 48.05 | 124.10 | -13.73 |
| IR | LQ | 80.4 | 0 | - | - | - | - | - | 15.75 | 0.75 | 55.71 | 125.17 | - |
|  | LQL | 1.0* | 0 | - | 0.51 | - | - | - | 24.72 | 0.78 | 55.40 | 128.39 | -3.22 |
|  | LQ<sub>ST</sub> | 84.2 | 0.09 | 0.1 | - | 1.50 | 7E-7 | - | 15.13 | 0.69 | 55.40 | 132.81 | -7.63 |
|  | LQ<sub>SD</sub> | 77.6 | 0 | - | - | - | - | 1.00 | 15.90 | 0.75 | 55.71 | 129.02 | -3.85 |
|  | LQL<sub>SD</sub> | 1.0* | 0 | - | 0.51 | - | - | 0.00* | 24.84 | 0.78 | 55.40 | 132.81 | -7.64 |
|  | LQ<sub>ST,SD</sub> | 65.1 | 0.16 | 0 | - | 1.47 | 8e-7 | 0.724 | 17.00 | 0.80 | 55.02 | 137.13 | -11.95 |

**Figure 1:**
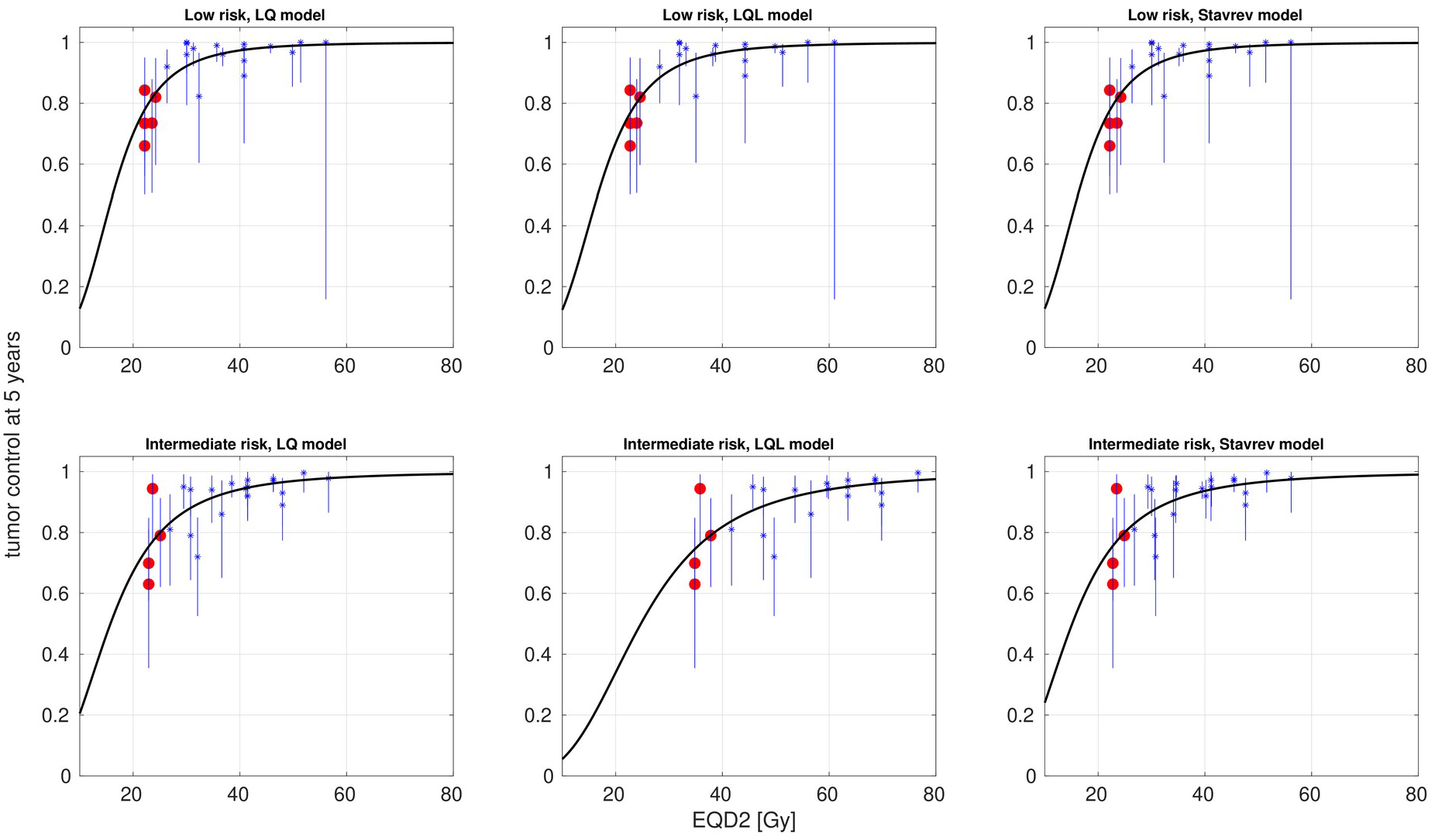
Best fits of LQ, LQL, and Stavrev’s reoxygenation models to dose–response data for prostate cancer treated with HDR-BT (low risk, top panels; intermediate risk, bottom panels). The *α*/*β* ratio was allowed to lie in a large interval (1 ≤ *α*/*β* ≤ 100 Gy). Clinical data (*) and 95% confidence intervals (bars), and modeled curves (solid lines). Single fraction schedules are highlighted as red circles.

In Table 3, we present the best-fitting parameters and the goodness-of-fit for the same models fitted to the same data, but forcing the *α*/*β* ratio to be low (1 ≤ *α*/*β* ≤ 8 Gy), as many radiobiological studies in external radiotherapy support a low *α*/*β* ratio. In Table 4 we report the 95% confidence intervals of the best-fitting parameters for the latter scenario. The analysis of the confidence intervals, being computationally demanding, was limited to the LQ model with and without incomplete repair correction and the LQL model. In Figure 2 we present TCP versus EQD2 curves (experimental data and model best-fits) for the LQ, LQL, and reoxygenation models without incomplete repair correction.

**Table 3:** Best fits obtained with the LQ, LQL, and Stavrev’s reoxygenation model, LQ_ST_, without or with (subscript SD) sublethal damage incomplete repair correction, separated by risk (low, LR, and intermediate risk, IR). The values of *α/β* were constrained to 1 ≤ *α*/*β* ≤ 8 Gy, to take into account the low *α/β* values typically reported for prostate cancer. The table shows best fitting parameters, maximum likelihood, AIC_c_, and ΔAIC_c_ (referred to the LQ model) values. The symbol * indicates that the best-fitting parameter reached the edge of the constraint window.

| Risk | Model | Parameters |  |  |  |  |  |  |  |  |  |  |  |
| --- | --- | --- | --- | --- | --- | --- | --- | --- | --- | --- | --- | --- | --- |
| | | $\alpha/\beta$<br>[Gy] | $\lambda'$<br>[Gy day<sup>-1</sup>] | $T_k$<br>[day] | $\delta$<br>[Gy<sup>-1</sup>] | $\varphi$ | $b$<br>[h<sup>-2</sup>] | $T_{repair}$<br>[h] | $D_{50}$<br>[Gy] | $\gamma_{50}$ | $-\log(L)$ | AIC<sub>c</sub> | ΔAIC<sub>c</sub> |
| LR | LQ | 8.0* | 1.00 | 35.54 | - | - | - | - | 41.0 | 1.84 | 60.21 | 134.43 | - |
|  | LQL | 1.0* | 1.33 | 31.45 | 0.48 | - | - | - | 27.2 | 1.09 | 47.23 | 112.45 | 21.98 |
|  | LQ<sub>ST</sub> | 8.0* | 0.60 | 28.93 | - | 8.5E-3 | 5.4E-5 | - | 41.0 | 1.84 | 60.15 | 142.91 | -8.48 |
|  | LQ<sub>SD</sub> | 8.0* | 0.00 | - | - | - | - | 2.75 | 42.8 | 1.72 | 55.29 | 128.59 | 5.84 |
|  | LQL<sub>SD</sub> | 1.0* | 1.33 | 31.45 | 0.48 | - | - | 0.00* | 27.2 | 1.09 | 47.23 | 117.07 | 17.36 |
|  | LQ<sub>ST,SD</sub> | 8.0* | 0.98 | 37.78 | - | 0.14 | 3.7E-5 | 3.62 | 40.9 | 1.33 | 54.34 | 136.67 | -2.24 |
| IR | LQ | 8.0* | 0.00* | - | - | - | - | - | 39.0 | 1.43 | 66.67 | 147.10 | - |
|  | LQL | 1.0* | 0.00* | - | 0.51 | - | - | - | 24.6 | 0.78 | 55.40 | 128.39 | 18.71 |
|  | LQ<sub>ST</sub> | 8.0* | 0.00* | - | - | 0.00* | 0.00* | - | 39.0 | 1.43 | 66.67 | 155.39 | -8.30 |
|  | LQ<sub>SD</sub> | 8.0* | 0.00* | - | - | - | - | 3.00 | 41.0 | 1.34 | 61.59 | 140.78 | 6.31 |
|  | LQL<sub>SD</sub> | 1.0* | 0.00* | - | 0.51 | - | - | 0.00* | 24.7 | 0.78 | 55.40 | 132.79 | 14.30 |
|  | LQ<sub>ST,SD</sub> | 8.0* | 0.00* | - | - | 0.00* | 0.00* | 3.00 | 41.0 | 1.34 | 61.59 | 150.26 | -3.16 |

**Table 4:** 95% confidence intervals of best fitting parameters for the LQ and LQL models without incomplete repair correction, and LQ model with incomplete repair correction (LQ_SD_). Results are separated by risk, low (LR) and intermediate (IR). The symbol * indicates that the parameter value reached the edge of the constraint window.

| Risk | Model | Parameters |  |  |  |  |  |  |
| --- | --- | --- | --- | --- | --- | --- | --- | --- |
| | | $\alpha/\beta$ [Gy] | $\lambda'$ [Gy day<sup>-1</sup>] | $T_k$ [day] | $\delta$ [Gy<sup>-1</sup>] | $T_{repair}$ [h] | $D_{50}$ [Gy] | $\gamma_{50}$ |
| LR | LQ | [7.1, 8*] | [0*, 2*] | [0*, 42*] | - | - | [36.0, 44.4] | [1.4, 2.3] |
|  | LQL | [1*, 8*] | [0*, 2*] | [0*, 42*] | [0.10, 1*] | - | [12.3, 48.7] | [0.7, 1.6] |
|  | LQ<sub>SD</sub> | [6.6, 8*] | [0*, 2*] | [0*, 42*] | - | [1.6, 4.9] | [36.0, 47.3] | [1.2, 2.1] |
| IR | LQ | [7.4, 8*] | [0*, 2*] | [0*, 42*] | - | - | [32.1, 43.2] | [1.0, 1.9] |
|  | LQL | [1*, 8*] | [0*, 2*] | [0*, 42*] | [0.06, 1*] | - | [10.2, 44.4] | [0.5, 1.3] |
|  | LQ<sub>SD</sub> | [6.8, 8*] | [0*, 2*] | [0*, 42*] | - | [1.6, 6*] | [31.5, 45.8] | [0.7, 1.8] |

**Figure 2:**
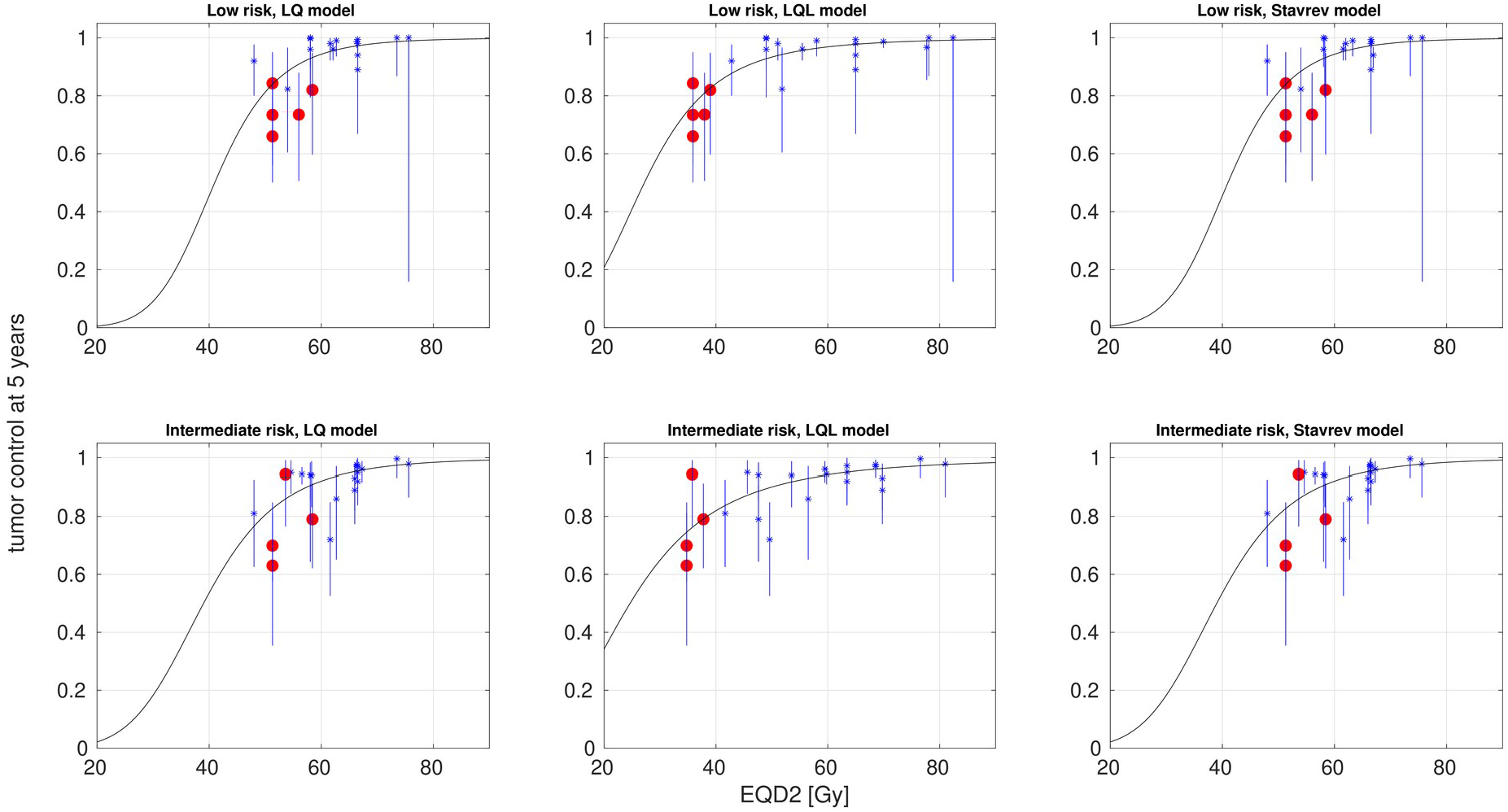
Best fits of LQ, LQL, and Stavrev’s reoxygenation models to dose–response data for prostate cancer treated with HDR-BT (low risk, top panels; intermediate risk, bottom panels). The *α*/*β* ratio was constrained to be low (1 ≤ *α*/*β* ≤ 8 Gy). Clinical data (*) and 95% confidence intervals (bars), and modeled curves (solid lines). Single fraction schedules are highlighted as red circles.

## Discussion

The response of prostate cancer to radiotherapy has been extensively studied, and the consensus is that the *α*/*β* ratio of prostate cancer is low **[7-9, 13, 14]**. This makes this tumor very sensitive to fractionation. Many external radiotherapy hypofractionated protocols have been investigated for prostate cancer **[11]**. Several studies have suggested that the LQ model may fail to describe tumor response at large doses per fraction **[16-18]** (even though the why and the how are not entirely clear, with different studies suggesting that the LQ may underestimate/overestimate the damage at large doses per fraction). In fact, recent analyses of the response of prostate cancer to SBRT reported a slight moderation of the LQ-predicted response at large doses **[13, 14]**.

The possible moderation of the damage with increasing dose is soft at the doses employed for SBRT, and clinical trials of SBRT for prostate cancer still reported high tumor control with doses per fraction up to 10 Gy **[14]**. This is not the case in HDR-BT, which is delivered with protocols that are more radical than those of SBRT, reaching >20 Gy in a single fraction. This makes HDR-BT an ideal scenario to investigate dose-response at large doses per fraction and the potential failure of the LQ model at such doses. Several HDR-BT clinical trials have shown a marked reduction in tumor control when delivering ∼20 Gy in a single fraction **[21–25]**, including very recent studies **[46]** (not analyzed in this work). This important loss in tumor control is not supported by a low *α*/*β* ratio and an LQ behaviour at large doses: for example, assuming *α*/*β=*3 Gy and using the LQ model, a 20 Gy × 1 treatment would be roughly isoeffective to 8 Gy × 5, and more effective than a conventional 2 Gy × 37 (BEDs of 153.3, 146.7, and 123.3 Gy_3_, respectively, ignoring proliferation). This led Guirado *et al*. to suggest a large *α*/*β* ratio in a recent analysis of tumor response to HDR-BT **[15]**. However, such a large *α*/*β* ratio is not consistent with many radiobiological studies that found a low *α*/*β* ratio for prostate cancer. It may be that the LQ model is indeed not adequate to describe the response to very large doses per fraction **[16]**.

In this study, we investigated the dose–response of prostate cancer to HDR-BT from a dataset containing 21 schedules (1643 patients) for LR and 22 schedules (1615 patients) for IR, with doses per fraction ranging from 6 to 20.5 Gy per fraction (Table 1, Supplementary Table 1). Our analysis specifically focused on investigating the LQ and alternative models to characterize dose-response at such large doses. Because the clinical data point to an overestimation of cell killing effect by the LQ at large doses, we investigated the LQL model **[19]**, which includes a moderation of cell killing at large doses. This particular response at large doses could also be caused by the role of hypoxia/reoxygenation, as originally suggested by Nahum *et al*. **[10]**. If re-oxygenation plays an important role in the response to fractionated radiotherapy, extremely hypofractionated protocols might lose tumor control. To investigate the effect of reoxygenation, we have used the simple model proposed by Stavrev *et al*. **[41]**, which accounts for reoxygenation through time-dependent *α* and *β* parameters. Because several schedules in the dataset delivered multiple fractions per day, we also investigated the role of incomplete repair on the modeling of response to treatment in each of the three models under investigation.

We followed two strategies for data fitting. First, we imposed broad constraints on the values of the best-fitting parameters. When following this strategy, the LQ proved superior to both the LQL and reoxygenation models to describe dose-response, with ΔAIC_c_^LQL^=-2 and ΔAIC_c_^Sta^=-8 (Table 2, Figure 1). However, in order to fit the data, the LQ required a large *α*/*β* value (95% confidence intervals 28-100 Gy and 17-100 Gy for LR and IR, respectively; Supplementary Table 2). This agrees with **[15]**, who found a large *α*/*β* ratio (∼23 Gy) from the analysis of a smaller dataset. The inclusion of incomplete repair in the modeling of response to treatment does not improve data fitting. For prostate cancer, studies have shown sublethal repair rates characterized by half-lifes *T*_*repair*_ ∼1.5-2 h **[47]**. The effect of such a repair rate would be small for times between fractions ≥6 h, the typical time between fractions in the studies analyzed in this work.

The large *α*/*β* ratio obtained from this fitting strategy is not in agreement with many studies analyzing dose-response of prostate cancer treated with external radiotherapy. Therefore, we investigated a second fitting strategy where the value *α*/*β* ratio was constrained to be low, 1 ≤ *α*/*β* ≤ 8 Gy. These limits were qualitatively set to double the 95% confidence intervals reported in Ref. **[14]**. When forcing the *α*/*β* to be low, the results were quite different (Table 3, Figure 2), and the LQL model became superior to both the LQ and reoxygenation models, ΔAIC_c_ ^LQL^∼22 for LR and 19 for IR. Analyses based on the AIC typically demand ΔAIC_c_>10 to state the superiority of a given model **[48]**. Using the best fitting parameters of the LQL (*α*/*β*=1 Gy, *δ*=0.48 Gy^-1^, for LR) the BED calculated with the LQL model for a 20 Gy × 1 treatment (95 Gy_1_) is far less than that of a conventional 2 Gy × 37 fractionation (184 Gy_1_).

Interestingly, while the superiority of the LQL over the LQ is clear, the re-oxygenation model does not improve the performance of the LQ model. This cannot be used to conclude that reoxygenation does not play a role in the response to HDR-BT, and may simply be due to the simplicity of the model considered. In particular, the implementation of time variation of *α* and *β* due to reoxygenation is not dose/treatment dependent. The study of other more complex models accounting for hypoxia and reoxygenation has not been addressed in this work. For example, the models proposed by Kuperman & Lubich **[49]** or Jeong *et al*. **[50]**.

The possibility that the LQ model overestimates the effect on prostate cancer of large doses per fraction has already been discussed in the context of external radiotherapy. Refs. **[13, 14]** found some evidence of a moderation of the LQ predicted effect with increasing doses. HDR-BT is delivered with more radical protocols than SBRT, >20 Gy in a single fraction, therefore the moderation of the effect can be more significant. Such radical hypofractionations are currently under investigation in external radiotherapy, with Zilli *et al*. currently investigating a 19 Gy × 1 fractionation [51]. The results of this and future clinical trials will shed more light on the response of prostate cancer to extreme hypofractionation.

In this study we have focused on studying potential radiobiological reasons for the decline of tumor control in extremely hypofractionated HDR-BT. A potential dosimetric origin of such low control (target coverage, dose homogeneity) was not investigated. Recently, Kuperman & Lubich **[52]** modeled the effect of target dose heterogeneities on the BED, and found that dose heterogeneities reduce the BED compared to the homogeneous dose scenario, especially for hypofractionated schedules. This effect might also explain the origin of the observed loss of tumor control for extreme hypofractionation, and merits further investigation.

Our study presents some limitations. In particular, the limited number of patients/schedules and the heterogeneity of the dataset may increase the uncertainties and potential sources of bias of the analysis by including different studies that may use different margins, different dose constraints, different dose calculation algorithms, etc. Another limitation was that we only analyzed a limited number of dose–response models, as discussed above.

## Conclusions

This analysis showed that the dose–response curves of prostate cancer to hypofractionated HDR-BT (LR and IR) is well described by the LQ model if the *α*/*β* is large (∼90 Gy), far beyond the values reported in several radiobiological studies of dose–response of prostate cancer treated with external radiotherapy. If the *α*/*β* is constrained to be low (≤8 Gy), the LQ model cannot fit the dose–response curves and the LQL proves the superior model. This is in agreement with recent studies in external radiotherapy that have found evidence of a moderation of the LQ-predicted effect with increasing dose per fraction. This moderation of the effect with increasing dose per fraction might affect dose and/fractionation prescription for prostate cancer.

The origin of the loss of control of radical single-fraction HDR-BT treatments merits further investigation: while in this work a reoxygenation model did not fit the data as well as the LQL model, more complex reoxygenation models might provide better fits to the clinical data; also, target dose heterogeneity may lead to patterns like those observed experimentally (loss of effectiveness for extreme hypofractionations), and should be further explored.

## Data Availability

All data produced in the present work are contained in the manuscript.

## Acknowledgements

This project has received funding from Ministerio de Ciencia e Innovación, Agencia Estatal de Investigación and FEDER, UE (grant PID2021-128984OB-I00), and Xunta de Galicia, Axencia Galega de Innovación (grant IN607D 2022/02).

