## Extended methods; supplementary table 1; supplementary table 2 for "Radiobiological meta-analysis of the response of prostate cancer to high dose rate brachytherapy"

*Supplementary Materials for "**Radiobiological meta-analysis of the response of prostate cancer to high dose rate brachytherapy**"*

### 1. Calculation of the EQD2 for different models

The *equivalent dose in 2 Gy fractions, EQD2,* of a given schedule is calculated by imposing iso-effectiveness with a 2 Gy/fraction treatment. There is some ambiguity in the calculation of the EQD2 regarding the effects that are included in the computation of the effectiveness of the 2 Gy/fraction treatment. Here, we will follow **[14]** and include *dose effects* (like the moderation of the quadratic term with increasing dose in the LQL model), but will ignore *time effects* (like incomplete repair or proliferation), which would require to assign a given schedule to the 2 Gy/fraction treatment (e.g. weekends off or not). The form of the EQD2 for each model can be calculated analytically as:

**LQ model**

$$EQD2_{LQ}=\frac{\left(D+\frac{dD}{(\alpha/\beta)}-\left(\frac{\lambda}{\alpha}\right)max\left(0,T-T_k\right)\right)}{\left(1+\frac{2}{(\alpha/\beta)}\right)}$$

If we include the effect of sublethal damage incomplete repair we obtain:

$$EQD2_{LQ,SD}=\frac{\left(D+\frac{dD}{(\alpha/\beta)}+\frac{2d^2\sum_{k=1}^{n}\left(\sum_{p=1}^{k-1}\left(\prod_{q=p}^{k-1}\theta_q\right)\right)}{(\alpha/\beta)}-\left(\frac{\lambda}{\alpha}\right)\max\left(0,T-T_k\right)\right)}{\left(1+\frac{2}{(\alpha/\beta)}\right)}$$

**LQL model**

$$EQD2_{LQL}=\frac{\left(D+\frac{2\left(\delta d+\exp\left(-\delta d\right)-1\right)D}{(\alpha/\beta)d\delta^2}-\left(\frac{\lambda}{\alpha}\right)max\left(0,T-T_k\right)\right)}{\left(1+\frac{2\delta+\exp\left(-2\delta\right)-1}{(\alpha/\beta)\delta^2}\right)}$$

If we include the effect of sublethal damage incomplete repair we obtain:

$$EQD2_{LQL,SD}=\frac{\left(D+\frac{dD\left(\delta d+\exp\left(-\delta d\right)-1\right)}{\left(\alpha/\beta\right)\left(\delta d\right)^2}+\frac{2d^2\left(\delta d+\exp\left(-\delta d\right)-1\right)\sum_{k=1}^{n}\left(\sum_{p=1}^{k-1}\left(\prod_{q=p}^{k-1}\theta_q\right)\right)}{\left(\alpha/\beta\right)\left(\delta d\right)^2}-\left(\frac{\lambda}{\alpha}\right)\max\left(0,T-T_k\right)\right)}{\left(1+\frac{\left(2\delta+\exp\left(\delta d\right)-1\right)}{\left(\alpha/\beta\right)\delta^2}\right)}$$

**Re-oxygenation model**

$$EQD2_S=\frac{\frac{1}{\alpha_0}\left(\sum_{i=1}^{n}\alpha_i d_i+\sum_{i=1}^{n}\beta_i d_i^2-\lambda max\left(0,T-T_k\right)\right)}{1+\frac{2}{\left(\alpha_0/\beta_0\right)}}$$

where $\alpha_i$ and $\beta_i$ refer to the values of $\alpha$ and $\beta$ at the time $t=t_i$ of delivery of the i-th fraction.

If we include the effect of sublethal damage incomplete repair we obtain:

$$EQD2_{S,SD}=\frac{\frac{1}{\alpha_0}\left(\sum_{i=1}^{n}\alpha_i d_i+\sum_{i=1}^{n}\beta_i d_i^2+2\sum_{i=1}^{n}\beta_i d_i\left(\sum_{p=1}^{i-1}d_p\left(\prod_{q=p}^{i-1}\theta_q\right)\right)-\lambda max\left(0,T-T_k\right)\right)}{1+\frac{2}{\left(\alpha_0/\beta_0\right)}}$$

### 2. Supplementary Tables

**Supplementary Table 1:** Detailed information of the analyzed schedules for low (LR) and intermediate risk (IR) prostate cancer, including: number of patients (N); dose per fraction (d); number of fractions (n); total dose (D); irradiation schedule derived from the publications, and presented as the time in hours at which each fraction is delivered (for modeling incomplete repair between fractions); overall treatment time (OTT, defined as treatment time - 1 day for modeling proliferation); percentage of patients receiving ADT, control at five years (TCP); and the first author and year of the study.

| Risk | N | d (Gy) | n | D (Gy) | Schedule (hours) | OTT (days) | ADT (%) | TCP (%) | Reference |
|---|---|---|---|---|---|---|---|---|---|
| LR | 288 | 7.25 | 6 | 43.5 | [0 6 24 168 174 192] | 8 | 0.0[*] | 98.7 | Hauswald (2015) |
| LR | 19 | 10.0 | 3 | 30.0 | [0 6 24] | 1 | 26.3 | 82.3 | Barkati (2012) |
| LR | 198 | 11.5 | 3 | 34.5 | [0 504 1008] | 42 | 5.0 | 96.1 | Strouthos (2017) |
| LR | 47[a] | 15.0 | 3 | 45.0 | [0 480 984] | 41[e] | 87.0 | 96.7 | Kukielka (2015) |
| LR | 233 | 9.5 | 4 | 38.0 | [0 6 24 30] | 1 | 0.0[**] | 98.0 | Jawad (2015) |
| LR | 48 | 12.0 | 2 | 24.0 | [0 6] | 0 | 0.0[***] | 92.0 | Jawad (2015) |
| LR | 56 | 13.5 | 2 | 27.0 | [0 6] | 0 | 0.0[+] | 100.0 | Jawad (2015) |
| LR | 44 | 19.0 | 1 | 19.0 | [0] | 0 | 34.0 | 66.0 | Prada (2016) |
| LR | 103 | 9.5 | 4 | 38.0 | [0 24 30 48] | 2 | 0.0[++] | 99.4 | Behmueller (2021) |
| LR | 19 | 9.5 | 4 | 38.0 | [0 6 24 30] | 1 | 0.0 | 89.0 | Johansson (2021) |
| LR | 85 | 11.0 | 3 | 33.0 | [0 336 672] | 27 | 0.0 | 99.0 | Johansson (2021) |
| LR | 69 | 14.0 | 2 | 28.0 | [0 336] | 13 | 0.0 | 98.0 | Johansson (2021) |
| LR | 23 | 19.0 | 1 | 19.0 | [0] | 0 | 0.0 | 84.3 | Morton (2020) |
| LR | 16 | 13.5 | 2 | 27.0 | [0 168] | 7 | 0.0 | 99.7 | Morton (2020) |
| LR | 196 | 9.5 | 4 | 38.0 | [0 6 336 342] | 13 | 3.6 | 94.0 | Tselis (2013) |
| LR | 84[b] | 13.5 | 2 | 27.0 | [0 6] | 0 | 32.7 | 96.0 | Nagore (2018) |
| LR | 26 | 7.0 | 7 | 49.0 | [0 6 24 30 48 54 72] | 3 | 7.7 | 100.0 | Yamazaki (2018) |
| LR | 2 | 6.0 | 9 | 54.0 | [0 6 24 30 48 54 72 78 96] | 4 | 0.0 | 100.0 | Yamazaki (2018) |
| LR | 22 | 20.5 | 1 | 20.5 | [0] | 0 | 68.2 | 82.0 | Prada (2018) |
| LR | 25[c] | 20.0 | 1 | 20.0 | [0] | 0 | 0.0 | 73.5 | Levi (2022) |
| LR | 40[d] | 19.0 | 1 | 19.0 | [0] | 0 | 0.0 | 73.4 | Siddiqui (2019) |

[*]42 patients out of 448 (288 LR and 160 IR) received ADT. Because ADT is most likely prescribed to HR/IR patients, we assumed that 0/288 LR patients received ADT.

[**] 61 patients out of 319 (233 LR, 86 IR) received ADT. Because ADT is most likely prescribed to HR/IR patients, we assumed that 0/233 LR patients received ADT.

[***] 4 patients out of 79 (48 LR, 31 IR) received ADT. Because ADT is most likely prescribed to HR/IR patients, we assumed that 0/48 LR patients received ADT.

[+]3 patients out of 96 (56 LR, 40 IR) received ADT. Because ADT is most likely prescribed to HR/IR patients, we assumed that 0/96 LR patients received ADT.

[++]33 patients out of 141 (103 LR, 32 IR, 6 HR) received ADT. Because ADT is most likely prescribed to HR/IR patients, we assumed that 0/103 LR patients received ADT.

[a]47 out of 77 patients were LR (61%). Biochemical control (BC) was not specified by risk group, and we assigned the overall BC (96.7%) to the LR group.

[b] 84 out of 119 patients were LR (71%). Biochemical control (BC) was not specified by risk group, and we assigned the overall BC (96.0%) to the LR group.

[c] 25 out of 33 were LR (76%). Biochemical control (BC) was not specified by risk group, and we assigned the overall BC (73.5%) to the LR group.

[d]40 out of 68 patients were LR (59%). Biochemical control (BC) was not specified by risk group, but the study found "No significant difference between low- and intermediate-risk patients". We assigned the overall BC (73.5%) to the LR group.

[e] median value

| | | | | | | | | | |
|---|---|---|---|---|---|---|---|---|---|
| IR | 54[*] | 6.50 | 7 | 45.5 | [0 6 24 30 48 54 72] | 3 | 44.3 | 93.0 | Yoshioka (2016) |
| IR | 160 | 7.25 | 6 | 43.5 | [0 6 24 168 174 192] | 8 | 26.3 | 97.6 | Hauswald (2015) |
| IR | 284 | 6.50 | 6 | 39.0 | [0 5 24 408 413 432] | 18[+] | 16.2 | 94.4 | Rogers (2015) |
| IR | 190[**] | 7.25 | 6 | 43.5 | [0 6 24 168 174 192] | 8 | 0.0 | 97.0 | Patel (2016) |
| IR | 135 | 11.50 | 3 | 34.5 | [0 504 1008] | 42 | 11.9 | 96.1 | Strouthos (2017) |
| IR | 86 | 9.50 | 4 | 38.0 | [0 6 24 30] | 1 | 70.9 | 95.0 | Jawad (2015) |
| IR | 31 | 12.00 | 2 | 24.0 | [0 6] | 0 | 12.9 | 81.0 | Jawad (2015) |
| IR | 40 | 13.50 | 2 | 27.0 | [0 6] | 0 | 7.5 | 79.0 | Jawad (2015) |
| IR | 28 | 19.50 | 1 | 19.5 | [0] | 0 | 53.6 | 94.4 | Hoskin (2017) |
| IR | 69 | 13.00 | 2 | 26.0 | [0 6] | 0 | 52.2 | 95.0 | Hoskin (2017) |
| IR | 49 | 10.50 | 3 | 31.5 | [0 6 24] | 1 | 71.0 | 94.0 | Hoskin (2017) |
| IR | 32 | 9.50 | 4 | 38.0 | [0 24 30 48] | 2 | 84.4 | 97.2 | Behmueller (2021) |
| IR | 22 | 11.00 | 3 | 33.0 | [0 336 672] | 27 | 0.0 | 86.0 | Johansson (2021) |
| IR | 34 | 14.00 | 2 | 28.0 | [0 336] | 13 | 0.0 | 72.0 | Johansson (2021) |
| IR | 64 | 19.00 | 1 | 19.0 | [0] | 0 | 0.0 | 69.9[***] | Morton (2020) |
| IR | 67 | 13.50 | 2 | 27.0 | [0 168] | 7 | 0.0 | 94.1[***] | Morton (2020) |
| IR | 81 | 9.50 | 4 | 38.0 | [0 6 336 342] | 13 | 23.5 | 92.0 | Tselis (2013) |
| IR | 48 | 6.50 | 7 | 45.5 | [0 6 24 30 48 54 72] | 3 | 22.9 | 89.0 | Yamakazi (2018) |
| IR | 52 | 7.00 | 7 | 49.0 | [0 6 24 30 48 54 72] | 3 | 96.2 | 99.6 | Yamakazi (2018) |
| IR | 39 | 6.00 | 9 | 54.0 | [0 6 24 30 48 54 72 78 96] | 4 | 76.9 | 97.8 | Yamakazi (2018) |
| IR | 34 | 20.50 | 1 | 20.5 | [0] | 0 | 14.7 | 79.0 | Prada (2018) |
| IR | 16 | 19.00 | 1 | 19.0 | [0] | 0 | 31.3 | 63.0 | Prada (2016) |

[*]62% of the 79 patients were treated with this treatment plan. Other fractionations were employed, but since BC is not specified separately, we assigned the overall BC (93%) to this fractionation.

[**]83% of the patients received this treatment plan. Other fractionations were employed, but since BC is not specified separately, we assigned the overall BC (93%) to this fractionation.

[***] weighted average of the BC for IR favourable group and IR unfavourable group.

[+] mean value

**Supplementary Table 2:** 95% confidence intervals of best fitting parameters ($\alpha/\beta$, $\delta$) for the LQ and LQL models without incomplete repair correction. Results are separated by risk, low (LR) and intermediate (IR). The values of $\alpha/\beta$ were not constrained to be low ($1 \leq \alpha/\beta \leq 100$ Gy). The symbol * indicates that the parameter value reached the edge of the constraint window.

| Risk | Model | Parameters | |
|---|---|---|---|
| | | $\alpha/\beta$ [Gy] | $\delta$ [Gy$^{-1}$] |
| LR | LQ | [27.8, 100*] | - |
| | LQL | [1*, 100*] | [0*, 1*] |
| IR | LQ | [16.6, 100*] | - |
| | LQL | [1*, 100*] | [0*, 1*] |
